# Integrating Heart Rate Variability and Psychometric Assessments to Evaluate Stress and Burnout in ICU Nursing Staff: A Pilot Study

**DOI:** 10.1101/2024.09.05.24313119

**Authors:** Alberto Rubio-López, Teresa Sierra-Puerta, Alejandro Rubio Navas

## Abstract

**Background:** Intensive Care Units (ICUs) are recognized as highly demanding environments that significantly contribute to stress and burnout among nursing staff. Despite increasing concern over burnout in healthcare, the relationship between physiological stress indicators, such as heart rate variability (HRV), and psychometric assessments has not been thoroughly explored in this setting.

**Objective:** This pilot study aimed to evaluate the relationship between HRV metrics and psychometric assessments of stress and anxiety in ICU nursing staff. Additionally, it explored the influence of shift type, shift duration, demographic factors, and lifestyle habits on these stress indicators.

**Methods:** An observational cross-sectional pilot study was conducted with 24 ICU healthcare professionals at a University Hospital in Madrid, Spain. HRV data were collected under controlled conditions, with measurements taken at the beginning and end of shifts. Psychometric assessments were conducted using the State-Trait Anxiety Inventory (STAI), Perceived Stress Scale (PSS-14), Nursing Stress Scale (NSS), and a Visual Analogue Scale for Stress (VASS). Non-parametric statistical tests were used to analyze correlations between HRV metrics, psychometric scores, and demographic/lifestyle variables.

**Results:** Significant negative correlations were observed between HRV metrics and perceived stress levels, particularly between the LF/HF ratio and stress measures. Night and extended shifts were associated with elevated stress, as indicated by lower HRV and higher psychometric stress scores. These findings suggest that shift type and duration significantly influence stress levels in ICU nursing staff.

**Conclusion:** This pilot study highlights the potential of HRV as an objective measure of stress in ICU nursing staff, with significant correlations observed between HRV metrics and psychometric assessments. The findings suggest that HRV could be a valuable tool for monitoring stress in real-time and identifying individuals at risk of burnout. However, further research with larger samples and a longitudinal approach is needed to validate these results and explore their implications for occupational health practices in ICU settings.

## Introduction

Intensive Care Units (ICUs) represent some of the most demanding environments in healthcare, requiring constant vigilance, rapid decision-making, and the management of life-threatening situations. Healthcare professionals working in ICUs, particularly nurses and healthcare assistants, are frequently exposed to important levels of stress due to the need to care for critically ill patients, execute complex medical interventions, and respond to unpredictable emergencies(1). Over time, this chronic exposure to such high-stress conditions can lead to burnout—a syndrome characterized by emotional exhaustion, depersonalization, and a diminished sense of personal accomplishment(2).

Burnout has been extensively studied across various healthcare settings due to its significant implications. Research shows that burnout adversely affects the mental and physical well-being of healthcare providers and compromises the quality of patient care. In ICU settings, where patient outcomes are linked to the performance and mental state of the healthcare team, the consequences of burnout can be particularly severe(3). Studies have highlighted correlations between burnout and negative outcomes such as increased absenteeism, reduced job satisfaction, and higher turnover rates, all of which can impair clinical judgment and patient safety(4). Despite the substantial body of literature on burnout in healthcare, there remains a significant gap in understanding how objective physiological indicators of stress, such as heart rate variability (HRV), relate to psychometric assessments of stress among ICU personnel(5).

HRV is increasingly recognized as a reliable and non-invasive biomarker of autonomic nervous system (ANS) function, reflecting the balance between the sympathetic nervous system (SNS) and the parasympathetic nervous system (PNS)(6). The SNS is responsible for the body’s “fight or flight” response, while the PNS governs “rest and digest” activities. HRV measures the variation in time intervals between heartbeats, providing insights into autonomic regulation and overall physiological stress levels. A higher HRV typically indicates a healthy balance between the SNS and PNS, suggesting good autonomic flexibility and resilience to stress. Conversely, a lower HRV is associated with increased sympathetic activity, reduced parasympathetic activity, and consequently, higher levels of physiological stress. While HRV has been validated as a stress indicator in various populations, including athletes, military personnel, and general healthcare workers, research specifically focusing on HRV in ICU personnel is limited(7). Studies in other high-stress professions, such as law enforcement and emergency medical services, have demonstrated that HRV can effectively monitor stress and predict burnout, suggesting its potential applicability in ICU settings(8)

In addition to HRV, psychometric tools provide valuable insights into the subjective experience of stress. The State-Trait Anxiety Inventory (STAI) is widely used to assess both state and trait anxiety, offering a comprehensive evaluation of an individual’s anxiety levels(9). The Perceived Stress Scale (PSS-14) measures the extent to which participants perceive their lives as unpredictable, uncontrollable, and overloaded, providing a general assessment of perceived stress(10). The Nursing Stress Scale (NSS) is specifically designed to evaluate stress related to various aspects of the nursing profession, including workload, patient care, and interpersonal relationships. These tools have been validated in healthcare settings, including among Spanish-speaking populations, making them appropriate for use in this study(11). Previous research has shown that higher perceived stress levels, as measured by the PSS-14, are associated with lower HRV, indicating greater physiological stress. However, few studies have integrated these approaches in ICU settings, where stress levels are exceptionally high. This study aims to bridge this gap by combining HRV analysis with psychometric assessments to provide a comprehensive evaluation of stress in ICU nursing staff.

Shift work is another well-documented stressor in healthcare, particularly in ICUs where patient care is required around the clock. Nurses and healthcare assistants working night shifts or extended hours often report higher levels of stress and burnout compared to those on regular day shifts(12). The disruption of circadian rhythms, lack of adequate rest, and social isolation associated with night shifts contribute to this increased stress burden. Additionally, extended shifts can lead to fatigue, impairing cognitive function and decision-making abilities, which are critical in ICU settings. Studies have shown that night shift work is associated with lower HRV, indicating higher physiological stress, and an increased risk of burnout. Despite these findings, the specific impact of shift type and length on both HRV and psychometric stress measures in ICU settings has not been thoroughly explored.

Given the high prevalence of stress and burnout among ICU staff and the growing interest in HRV as a biomarker for stress, this study seeks to explore the utility of HRV in conjunction with psychometric assessments. By examining the correlations between HRV metrics and subjective stress measures across different shifts, this study aims to identify specific patterns and predictors of stress and burnout in this high-risk population. Additionally, the study will explore the influence of demographic and lifestyle factors on these stress responses, contributing to the broader literature on occupational stress and burnout in healthcare.

## Methods

### Study Design

This observational cross-sectional pilot study was conducted to explore the relationship between heart rate variability (HRV) metrics and psychometric assessments of stress in ICU nursing staff. The study took place over a three-month period at a University Hospital in Madrid, Spain (from February 5^th^ to April 26^th^ of 2024). Given the exploratory nature of this study and the small sample size, the primary goal was to identify potential correlations and trends that could inform future, larger-scale research. Due to the study’s cross-sectional design, the findings should be interpreted with caution, as no causal inferences can be made, and generalizability is limited. Future research could benefit from longitudinal designs and larger samples, potentially expanding the scope to other high-stress hospital settings, such as emergency rooms and operating theaters

### Participants

The original sample consisted of 26 ICU healthcare professionals; however, two participants were excluded from the analysis due to errors in signal acquisition and processing, resulting in a final sample size of twenty-four participants. This sample size was deemed appropriate for a pilot study aimed at identifying preliminary trends and correlations. Participants were recruited based on convenience sampling, with 54% of the ICU personnel volunteering to participate. The final sample included 62.5% registered nurses and 37.5% healthcare assistants (TCAEs)

### Inclusion Criteria

- Full-time employment in the ICU with at least one year of experience.
- Age between 25 and 50 years to minimize age-related variability in HRV, aligning with the actual demographic distribution of the ICU staff
- No history of cardiovascular disease or use of medications that could influence HRV (e.g., beta-blockers, antiarrhythmics).
- Willingness to participate, with all participants providing informed consent. The study was conducted in accordance with the principles outlined in the Declaration of Helsinki, ensuring strict adherence to ethical guidelines for medical research involving human subjects

### Exclusion Criteria

- Presence of any medical condition that could interfere with HRV readings or psychometric assessments.
- Recent surgery or trauma that could affect autonomic nervous system function.
- Errors in signal acquisition and processing with the Biosignalplux device, leading to the exclusion of two participants from the final analysis.

Participants were balanced in terms of gender and shift type (day vs. night) to ensure that the sample was representative of the broader ICU staff population. Demographic data, including age, gender, years of experience, and typical shift patterns, were collected through a structured questionnaire. A comprehensive lifestyle and health questionnaire was also administered to gather information on participants’ general health status, lifestyle habits, and any factors that could influence stress levels or HRV.

### Psychometric Assessments

To evaluate the subjective experience of stress and anxiety among participants, several validated psychometric instruments were employed:

- **State-Trait Anxiety Inventory (STAI)**: Measures both the current level of anxiety (state anxiety) and the general tendency to experience anxiety (trait anxiety), consisting of two subscales with twenty items each, scored on a four-point Likert scale.
- **Perceived Stress Scale (PSS-14)**: Assesses the extent to which participants perceive their lives as unpredictable, uncontrollable, and overloaded, with fourteen items scored from 0 (never) to 4 (very often).
- **Nursing Stress Scale (NSS)**: Specifically designed for nursing staff, this scale measures stress related to various aspects of the nursing profession, including workload, patient care, and interpersonal relationships, with thirty-four items scored on a five-point Likert scale.
- **Visual Analogue Scale for Stress (VASS)**: Administered at the beginning and end of the shift, allowing participants to rate their perceived stress on a continuum from 0 (no stress) to 100 (maximum stress).

These tools have been previously validated in Spanish-speaking populations, including healthcare settings, and were deemed appropriate for this study. Although there is an inherent risk of social desirability bias in self-reporting tools, participants were encouraged to provide honest responses to ensure the reliability of the data. The original Spanish versions of the STAI, PSS-14, and NSS questionnaires used in this study are provided as supplementary material to this paper (S1-Original Questionnaires).

### Biometric Measurements

HRV was the primary biometric indicator used to assess stress levels in this study. HRV data were collected using the Biosignalplux system, a portable ECG monitoring device that records real-time heart rate data and allows for detailed HRV analysis. The device was attached to participants using chest electrodes at the start and end of their shifts (Figure 1), with HRV measurements taken while participants were seated and at rest for 10 minutes before the start of the shift and another 10 minutes after the end of the shift (Figure 2). This standardized procedure ensured that the HRV data were collected in a controlled environment, minimizing potential confounding factors such as physical activity or movement.

**Figure 1:**
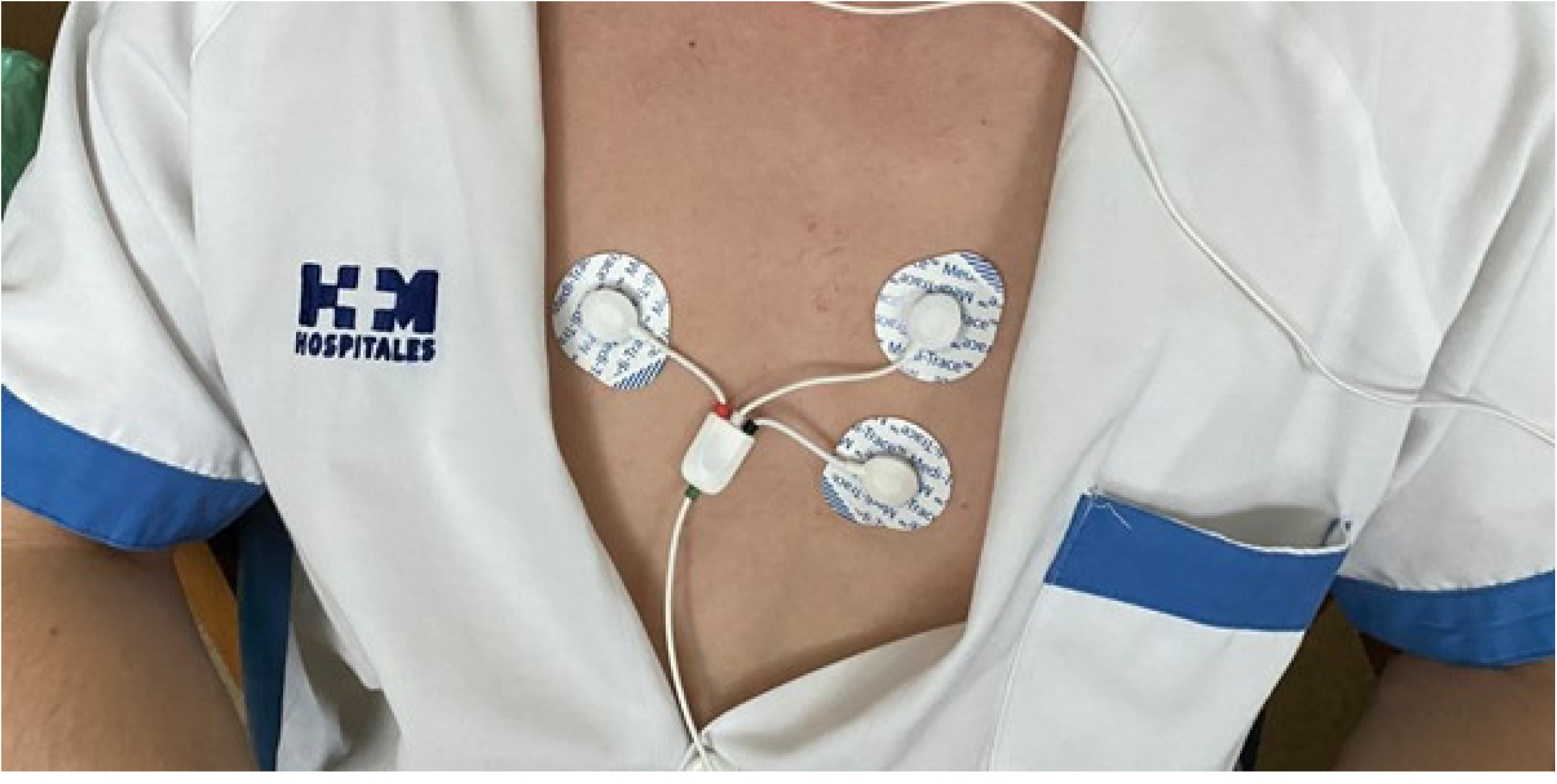
Electrode Placement for HRV Measurement: The image shows the placement of the Biosignalplux electrodes on a participant’s chest. Electrodes were placed according to the manufacturer’s guidelines to ensure accurate HRV data collection. The subject provided written consent for the use of this image in the publication

**Figure 2:**
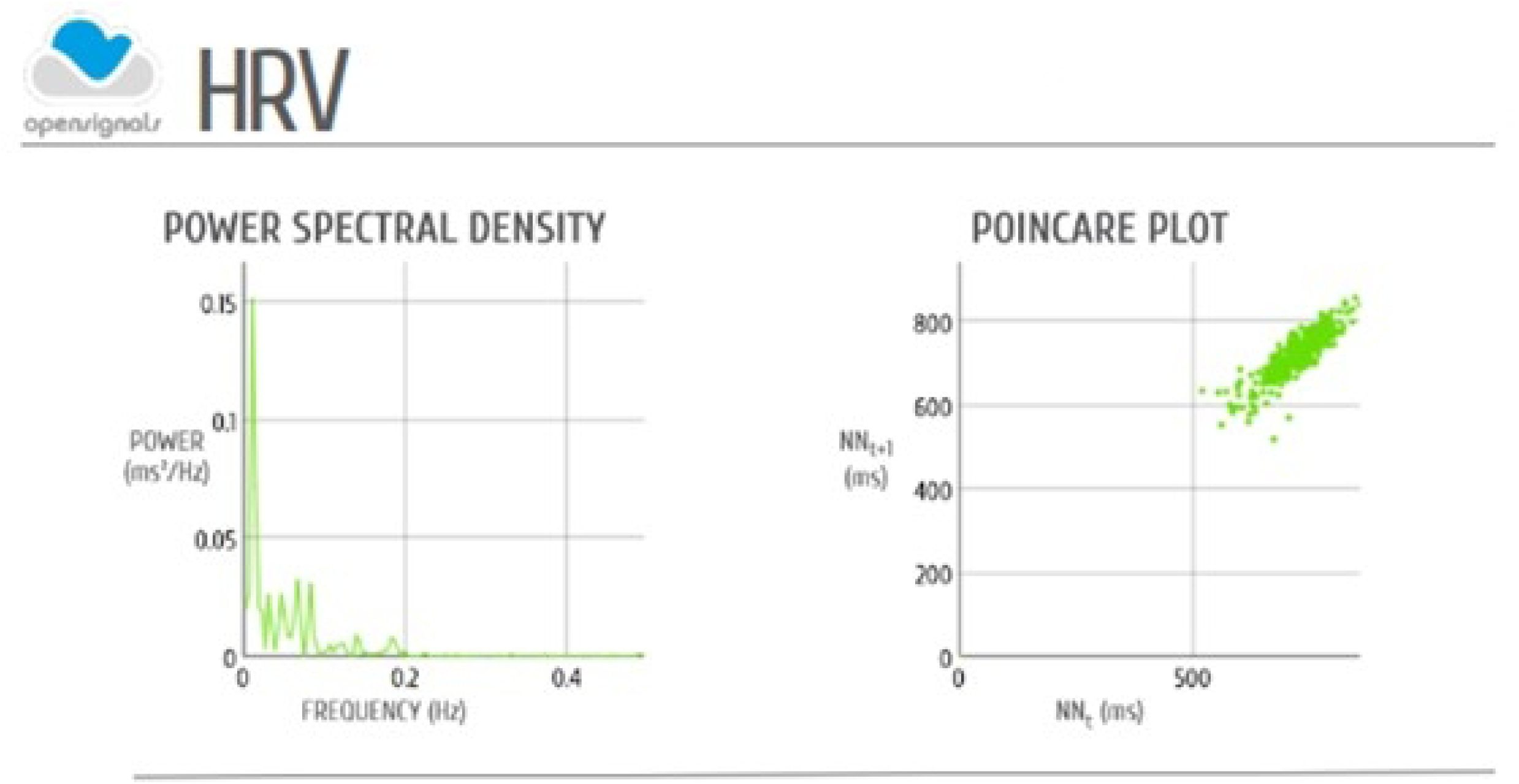
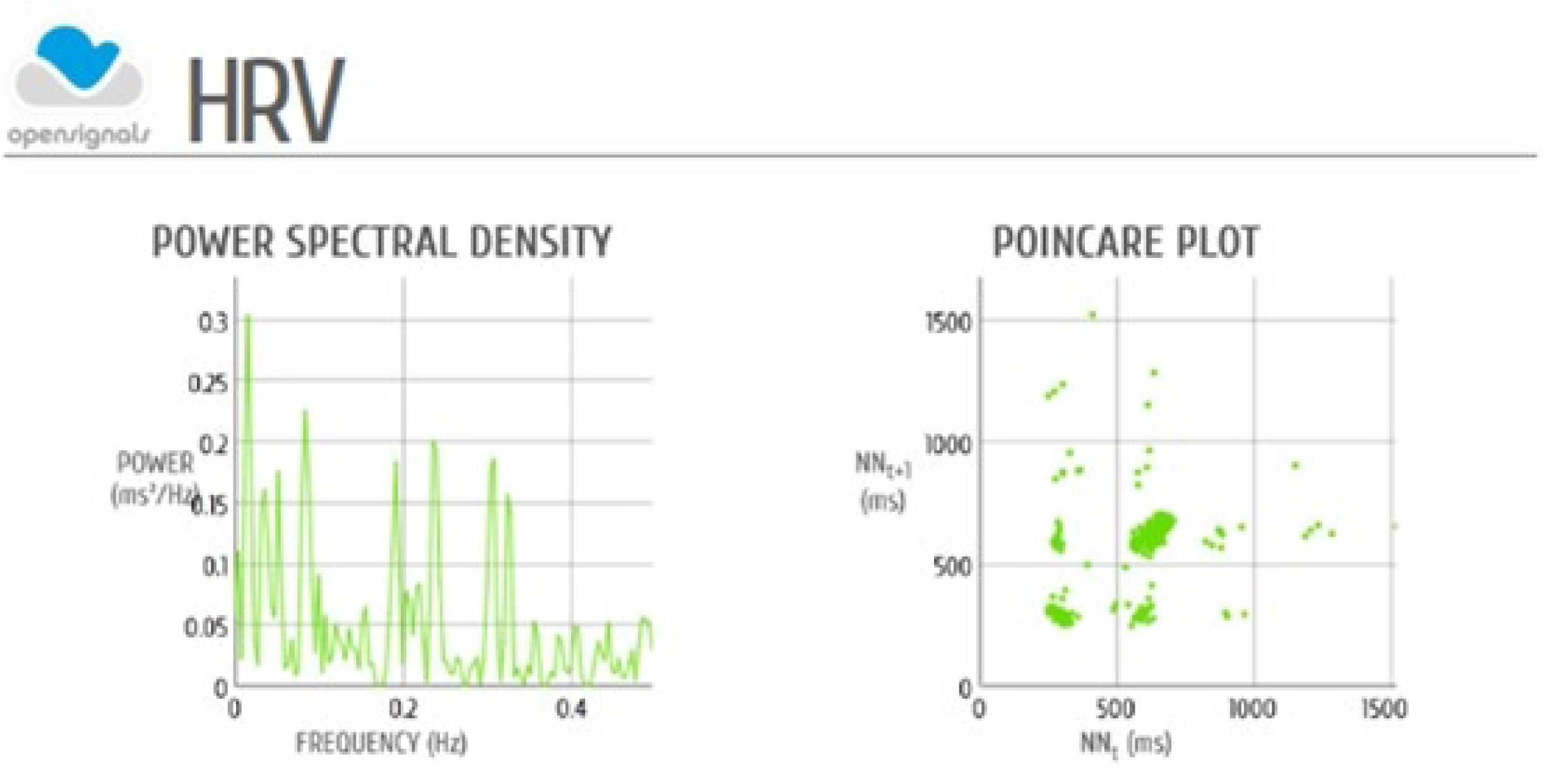
HRV Parameter Calculation Interface. The interface of the DSP software used to calculate HRV parameters such as RMSSD and LF/HF ratio at beginning (2.a) and end (2.b) of shift is shown

### Key HRV Parameters Analyzed

- **Root Mean Square of Successive Differences (RMSSD)**: A time-domain measure of HRV that reflects short-term variations in heart rate and is closely related to parasympathetic activity.
- **Percentage of Consecutive NN Intervals That Differ by More Than 50 ms (pNN50)**: Indicates the proportion of differences between adjacent heartbeats that exceed fifty milliseconds, associated with parasympathetic regulation.
- **Low-Frequency to High-Frequency Ratio (LF/HF Ratio)**: Represents the balance between sympathetic and parasympathetic nervous activity, with a higher ratio indicating increased sympathetic activity and reduced parasympathetic activity, often associated with stress.

### Secondary Biometric Measures

- **Blood Pressure**: Measured at the beginning, mid-point, and end of each shift to monitor potential cardiovascular responses to stress.
- **Respiratory Rate and Oxygen Saturation**: Monitored continuously alongside HRV to provide a comprehensive overview of physiological responses.

Electrode placement followed the manufacturer’s instructions, and participants were in a resting state during measurements to minimize potential movement artifacts. Secondary biometric measures were taken simultaneously with HRV data collection to ensure consistency.

### Procedure

Prior to data collection, participants received a detailed explanation of the study’s objectives and procedures. Informed consent was obtained, and participants were assured of the confidentiality of their data. Psychometric assessments and demographic questionnaires were completed at home before the beginning of the recording phase. Each participant was equipped with the Biosignalplux system at the beginning of their shift. The device was calibrated to ensure accurate data collection, and participants were instructed to continue their regular duties without altering their routines.

The study was designed to include both day and night shifts, with participants monitored during a minimum of two shifts. This approach allowed for a comparison of HRV and stress levels across different working conditions. HRV data were transmitted to a secure server for analysis immediately after each shift. Psychometric data were entered into a database, with each participant assigned a unique identifier to maintain anonymity. Data were anonymized and stored securely, with access limited to the principal investigator.

### Data Analysis

Statistical analysis was conducted using SPSS version 25.0. The primary analysis involved comparing HRV metrics across different shift types (day vs. night) and correlating these metrics with psychometric scores and demographic data. After the results were obtained and classified, a sequential statistical analysis was conducted. Initially, the normality of the samples was assessed using both the Kolmogorov-Smirnov and Shapiro-Wilk tests, considering the small sample size. Since several parameters did not follow a normal distribution and the sample size was limited, it was decided to use non-parametric tests for the analysis.

### Statistical Methods

- **Wilcoxon Signed-Rank Tests**: Used for comparative analysis of HRV and psychometric data at the start and end of shifts.
- **Spearman Correlations**: Employed to assess relationships between HRV metrics and psychometric scores.
- **Mann-Whitney U Tests**: Applied for comparative analysis between day and night shifts.
- **Multiple Regression Analysis (MANOVA)**: Conducted to explore interactions between demographic/medical variables and biometric/psychometric stress indicators, acknowledging that multicollinearity could pose challenges given the small sample size.

Significance was set at p < 0.05 for all analyses. Results were interpreted in the context of existing literature on HRV and occupational stress in healthcare professionals, with particular attention to the implications for managing stress in ICU settings. The pilot nature of the study and the small sample size necessitate caution in generalizing the findings, but they provide a basis for future research and potential interventions in similar high-stress environments.

## Results

### Demographic Characteristics

The study included twenty-four participants, of whom 62.5% were registered nurses, and 37.5% were healthcare assistants (TCAEs). The mean age of participants was 38 years, with 70.83% being female (Table 1). Most participants (50%) worked 12-hour shifts, and 54.17% were assigned to the morning shift. On average, participants had 3.8 years of experience in their current roles. The average household income was reported as €3,000, with nearly half of the participants indicating caregiving responsibilities for dependents.

**Table 1.** Demographic Characteristics.

| **Characteristic** | **Percentage (%)** |
| --- | --- |
| **Gender** |  |
| Female | 70.83 |
| Male | 29.17 |
| **Age** |  |
| < 30 years | 25.00 |
| 30-40 years | 45.83 |
| 40-50 years | 12.50 |
| > 50 years | 16.67 |
| **Household Income** |  |
| < €2500 | 33.34 |
| €2500-4500 | 54.16 |
| > €4500 | 12.50 |
| **Job Position** |  |
| Registered Nurse (DUE) | 62.50 |
| Healthcare Assistant (TCAE) | 37.50 |
| **Years of Service** |  |
| < 5 years | 66.67 |
| > 5 years | 33.33 |
| **Shift** |  |
| Morning | 54.17 |
| Afternoon | 12.50 |
| Night | 33.33 |
| **Shift Duration** |  |
| 7 hours | 29.17 |
| 10-12 hours | 70.83 |

Lifestyle habits were self-reported, with most participants consuming alcohol occasionally. Tobacco use was reported by 30% of participants, though less than 10% were significant smokers (Table 2). Additionally, 85% consumed 1-3 cups of coffee daily, and 90% engaged in regular physical activity. Notably, over half of the participants felt that stress negatively impacted their work performance, with 62.5% having considered leaving the profession and nearly 30% reporting having experienced suicidal ideation at some point (Table 3).

**Table 2.** Lifestyle and Consumption Habits.

| **Characteristic** | **Percentage (%)** |
| --- | --- |
| **Alcohol Consumption** |  |
| Never | 12.50 |
| Occasionally | 87.50 |
| **Tobacco Consumption** |  |
| Never | 70.83 |
| Regular (3+ cigarettes/day) | 16.66 |
| **Coffee Consumption** |  |
| None | 12.50 |
| 1-3 cups/day | 83.33 |
| **Recreational Drug Use** |  |
| Never | 87.50 |
| Occasionally | 12.50 |
| **Physical Activity** |  |
| None | 12.50 |
| Regular | 50.00 |
| **Perceived Stress Impact on Work** |  |
| Yes | 54.16 |
| **Considered Leaving Profession** |  |
| Yes | 62.50 |
| **Considered Self-Harm** |  |
| Yes | 29.16 |

**Table 3.**
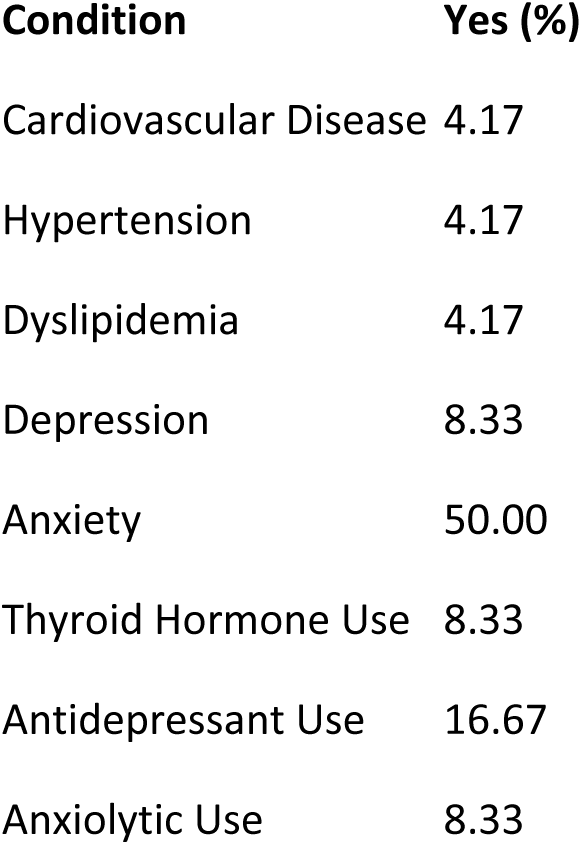
Health Status and Medication Use.

| Condition | Yes (%) |
| --- | --- |
| Cardiovascular Disease | 4.17 |
| Hypertension | 4.17 |
| Dyslipidemia | 4.17 |
| Depression | 8.33 |
| Anxiety | 50.00 |
| Thyroid Hormone Use | 8.33 |
| Antidepressant Use | 16.67 |
| Anxiolytic Use | 8.33 |

### Statistical Analysis and Normality Testing

Given the small sample size and the non-normal distribution of some parameters, non-parametric tests were applied throughout the analysis. This approach was deemed appropriate for identifying potential correlations in this pilot study, with the aim of informing larger-scale research.

### Basic Physiological Variables

- **Shift Duration Impact**: The Wilcoxon signed-rank test revealed that only the perceived stress scale (VASS) exhibited a statistically significant change (p < 0.05), increasing from the beginning to the end of the shift. This finding aligns with expectations that stress levels would escalate during the shift, consistent with previous research.
- **Shift Type Comparison**: The Kruskal-Wallis test showed no significant differences in most physiological variables (e.g., heart rate, oxygen saturation, systolic blood pressure) across different shifts (morning, afternoon, night). However, diastolic blood pressure demonstrated a significant variation (p = 0.028), suggesting that different shift types might exert varying levels of cardiovascular stress.
- **Work Shift Duration**: Comparisons between normal and extended shifts using the Kruskal-Wallis test revealed no statistically significant differences in the measured physiological variables, except for diastolic blood pressure, which is a well-documented indicator of stress levels in the literature.

### Biometric Variables

- **Shift Duration Impact**: Wilcoxon signed-rank tests indicated no significant differences in HRV metrics (RMSSD, pNN50, LF/HF ratio) between the beginning and end of shifts. This stability in HRV suggests that the physiological impact of stress, as measured by HRV, remained consistent throughout the work shift. However, this finding may be influenced by the small sample size, limiting the detection of subtler variations.
- **Shift Type Comparison**: The Kruskal-Wallis test revealed that most HRV parameters did not significantly differ between morning, afternoon, and night shifts. However, the LF/HF ratio at the beginning of the shift presented a significant difference across shifts (p = 0.043), indicating variations in autonomic nervous system activity depending on shift timing. This finding is consistent with the literature, which associates night shifts with increased sympathetic nervous system activity and higher stress levels.
- **Work Shift Duration**: Like the physiological variables, no significant differences were found in HRV metrics between normal and extended shifts, except for the LF/HF ratio, which showed a significant difference (p = 0.033). This suggests that longer shifts might contribute to an imbalance in autonomic regulation, particularly in the sympathetic-parasympathetic balance, which could be indicative of elevated stress levels.

### Psychometric Variables

The psychometric scales used in the study demonstrated high reliability, with a Cronbach’s alpha of 0.883 for the STAI scale (Table 4).

**Table 4.** Summary of Psychometric Scale Results. Psychometric scales and their respective score ranges: STAI – State (range 20-80), STAI – Trait (range 20-80), PSS (range 0-40), and NSS (higher scores indicate greater levels of job-related stress)

| ID | State Anxiety Score | Trait Anxiety Score | PSS Score | NSS Score |
| --- | --- | --- | --- | --- |
| 01 | 25 | 21 | 37 | 45 |
| 02 | 12 | 21 | 25 | 35 |
| 03 | 25 | 27 | 28 | 22 |
| 04 | 18 | 14 | 20 | 25 |
| 05 | 16 | 11 | 19 | 33 |
| 06 | 32 | 29 | 23 | 38 |
| 07 | 27 | 32 | 33 | 35 |
| 08 | 13 | 26 | 20 | 46 |
| 09 | 16 | 14 | 17 | 41 |
| 10 | 18 | 16 | 22 | 37 |
| 11 | 10 | 4 | 11 | 41 |
| 12 | 15 | 10 | 20 | 28 |
| 13 | 27 | 31 | 29 | 45 |
| 14 | 12 | 10 | 19 | 20 |
| 15 | 11 | 8 | 21 | 36 |
| 16 | 15 | 16 | 22 | 24 |
| 17 | 20 | 26 | 28 | 34 |
| 18 | 48 | 42 | 44 | 28 |
| 19 | 13 | 16 | 18 | 24 |
| 20 | 16 | 21 | 30 | 53 |
| 21 | 28 | 25 | 31 | 79 |
| 22 | 17 | 10 | 30 | 40 |
| 23 | 12 | 17 | 18 | 32 |
| 24 | 20 | 33 | 44 | 48 |

The following correlations were observed:

- **VASS and Anxiety**: Initial VASS scores were moderately correlated with state anxiety (STAI A_E) (r = 0.407, p < 0.05), and strong correlations were found between state anxiety (STAI A_E) and both trait anxiety (STAI A_R) (r = 0.793, p < 0.001) and perceived stress (PSS) (r = 0.703, p < 0.001). These correlations underscore the interrelationship between anxiety and perceived stress among ICU staff, which is consistent with findings from other studies in similar populations.
- **Shift Comparisons**: Multiple comparisons using the Kruskal-Wallis test, adjusted by Turkey’s HSD and Bonferroni corrections where applicable, indicated no significant differences in psychometric variables across different shifts or between normal and extended shifts. This lack of significant differences is due to the small sample size, which may have limited the statistical power to detect differences.

### Correlations Between Demographic Variables and Psychometric Tests

**Income and Anxiety**: Higher income levels were significantly correlated with lower levels of state anxiety (STAI_A_E: r = -0.443, p = 0.030) and trait anxiety (STAI_A_R: r = -0.489, p = 0.015). This finding suggests that a better economic status may mitigate anxiety levels among ICU staff.

### Lifestyle Factors

**Physical Activity**: There was a positive correlation between initial VASS scores and participation in sports (r = 0.422, p = 0.040), suggesting that individuals with higher initial stress may engage more in physical activities, potentially as a coping strategy.

### Health Status

**Medication Use**: The use of antidepressants was highly correlated with the use of anxiolytics (r = 0.674, p < 0.001), indicating that these medications are often co-prescribed for managing mental health conditions in this population.

### Self-Perceived Stress

**Anxiety and Stress**: Both state and trait anxiety were significantly correlated with perceived stress (PSS) and the initial VASS score. High levels of anxiety were also associated with lower well-being (STAI_A_E: r = -0.599, p = 0.002; STAI_A_R: r = -0.459, p = 0.024) and increased thoughts of “giving up” or “abandoning” (STAI_A_R: r = 0.723, p < 0.001). These findings highlight the significant impact of anxiety and stress on mental health, suggesting a potential area for targeted interventions.

### Global Correlations Between Biometric and Psychometric Variables

When analyzing the data globally across all time points (both start and end of shifts), a consistent pattern emerged where higher levels of psychological stress, as measured by validated psychometric scales, were associated with lower HRV metrics. This pattern supports HRV as a reliable indicator of autonomic dysregulation in response to stress. The high intercorrelation among HRV measures further reinforces their validity as robust indicators of cardiac autonomic function, consistent with findings in other studies of occupational stress in healthcare settings.

## Discussion

This study explored the relationships between biometric stress variables, derived from heart rate variability (HRV) analysis, and psychometric stress variables among ICU nursing staff. Additionally, the study examined the impact of shift type and duration on these variables and investigated the influence of demographic factors, health status, and lifestyle habits on stress and anxiety. The findings contribute to a growing body of literature on occupational stress in healthcare, with specific implications for ICU settings.

### HRV and Psychometric Stress Measures

A significant negative correlation was observed between HRV metrics, particularly the LF/HF ratio, and perceived stress levels, as measured by the Visual Analogue Scale for Stress (VASS). This finding aligns with previous research, such as the study by Xinxia Li et al., which demonstrated a significant association between HRV and perceived stress levels among nurses(13). The negative correlation suggests that higher perceived stress is associated with lower HRV, indicating increased physiological stress. These results support the use of HRV as a potential objective marker for stress in ICU nursing staff, providing a physiological correlation to subjective experiences of stress.

However, it is important to note that the stability of HRV metrics across shifts, with no significant changes observed between the beginning and end of shifts, might be influenced by the small sample size. This stability could suggest a level of physiological resilience among the participants, or it might reflect the limitations of detecting more subtle variations in autonomic regulation with the available sample(14). The lack of significant differences in HRV metrics between different shifts (morning, afternoon, night) further complicates the interpretation, although the significant variation in the LF/HF ratio across shifts points to potential differences in autonomic nervous system activity that warrant further investigation.

### Impact of Shift Work on Stress

The study found that shift type and duration significantly influenced certain stress indicators. Night shifts were associated with a higher LF/HF ratio at the beginning of the shift, indicating greater sympathetic nervous system activity and higher stress levels. This finding is consistent with existing literature that links night shifts to increased physiological stress and reduced HRV(15). For example, Varghese et al. found that night shift work was associated with lower HRV among healthcare workers, which is indicative of elevated stress.(16)

Extended shifts also exacerbated stress, as evidenced by the significant difference in diastolic blood pressure between normal and extended shifts(17). While HRV metrics did not show significant changes based on shift duration, the observed variations in blood pressure suggest that extended shifts may still pose a cardiovascular strain on ICU staff. This finding underscores the need for further research into the physiological impacts of extended work hours, especially in high-stress environments like ICUs.

### Psychometric Assessments and Their Correlations

Psychometric assessments revealed strong correlations between state and trait anxiety, perceived stress, and the initial VASS scores. These correlations indicate that individuals with higher baseline anxiety levels are more likely to report elevated stress throughout their shifts. This finding is consistent with studies such as those by Lesage et al. which demonstrated a significant association between anxiety symptoms and psychological stress among healthworkers(18). The persistence of anxiety and stress levels throughout the work shift suggests that these factors are deeply intertwined and may contribute to the overall burden of stress experienced by ICU staff.

### Demographic and Lifestyle Influences on Stress

Higher income levels were found to correlate significantly with lower levels of state and trait anxiety, suggesting that financial stability may serve as a buffer against stress and anxiety in this population. This is in line with the broader literature on socioeconomic status and mental health, where higher income often correlates with better coping mechanisms and lower perceived stress(19).

Additionally, physical activity was positively correlated with initial stress levels, indicating that those who engage in regular exercise might be using it as a coping strategy to manage higher baseline stress. This aligns with existing research suggesting that physical activity can serve as a protective factor against stress and burnout(20). The significant correlation between the use of antidepressants and anxiolytics further highlights the mental health challenges faced by ICU staff and suggests that medication use may be common among those experiencing elevated levels of stress and anxiety.(21)

### Implications for Practice

The findings of this study have several practical implications for managing stress and preventing burnout among ICU staff. The significant correlations between HRV and psychometric measures of stress suggest that HRV monitoring could be integrated into routine occupational health assessments to provide early warnings of stress-related health risks. By identifying shifts associated with increased stress, such as night shifts and extended work hours, healthcare administrators can implement targeted interventions, such as rotating shifts, reducing shift duration, and promoting physical activity as a coping strategy.

Furthermore, the strong association between anxiety, stress, and income levels suggests that financial incentives or support might play a role in mitigating stress. Programs aimed at improving the financial well-being of healthcare workers, alongside traditional stress management and mental health support services, could be beneficial.

### Limitations and Future Directions

This study has some limitations that should be addressed in future research. The small sample size is the most significant limitation, as it restricts the generalizability of the findings and may have prevented the detection of subtler differences in HRV metrics. The cross-sectional design also limits the ability to draw causal inferences between HRV, psychometric measures, and shift work. Future studies should include larger, more diverse samples and consider longitudinal designs to capture the progression of stress and burnout over time. Expanding the scope to include other high-stress hospital settings, such as emergency rooms and operating theaters, would also provide valuable comparative data.

Additionally, the study’s reliance on self-reported data for psychometric assessments introduces the possibility of social desirability bias, where participants might underreport their stress levels. To mitigate this, future research could incorporate more objective measures of stress, such as cortisol levels, alongside HRV and psychometric assessments.

## Conclusion

This pilot study highlights the potential of HRV as an objective measure of stress in ICU nursing staff, with significant correlations observed between HRV metrics and psychometric assessments. The findings suggest that HRV could be a valuable tool for monitoring stress in real-time and identifying individuals at risk of burnout. However, further research with larger samples and a longitudinal approach is needed to validate these results and explore their implications for occupational health practices in ICU settings. The integration of HRV monitoring with traditional stress management programs could lead to more effective interventions, improving the well-being and performance of ICU staff.

## Data Availability

The datasets used and/or analyzed during the current study are available in publicly available repositories: https://doi.org/10.6084/m9.figshare.26869255

## Acknowledgments

We sincerely thank all the nurses and healthcare assistants (TCAEs) who participated in this study. Your contributions were invaluable, and this research would not have been possible without your involvement. We appreciate your time and effort in helping us better understand the stress and burnout experienced in ICU settings

## DECLARATIONS

### Ethics approval and consent to participate

This study received ethical approval from the HM Hospitales Research Ethics Committee (Code: 18.12.1339.GHM) and involved twenty-six consenting ICU health personnel. All participants provided written informed consent, and their data were anonymized to protect confidentiality

### Competing interests

The authors declare that they have no competing interests.

### Funding

Not applicable

### Authors’ contributions

ARL designed the study, collected the data, performed the statistical analysis, and wrote the study as the principal investigator.

TSP participated in the performance tests and in the design and collection of biometric parameters, collected the data and participated writing of the paper.

ARN participated in the performance tests and in the design and collection of biometric parameters

## Notes

### Competing Interest Statement

The authors have declared no competing interest.

### Funding Statement

The author(s) received no specific funding for this work.

### Author Declarations

This study received ethical approval from the HM Hospitales Research Ethics Committee (Code: 18.12.1339.GHM)

